# Effectiveness of mRNA COVID-19 Vaccines among Employees in an American Healthcare System

**DOI:** 10.1101/2021.06.02.21258231

**Authors:** Nabin K. Shrestha, Amy S. Nowacki, Patrick C. Burke, Paul Terpeluk, Steven M. Gordon

## Abstract

**Background:** The mRNA SARS-CoV-2 vaccines have shown great promise in clinical trials. The purpose of this study was to evaluate the effectiveness of these vaccines under real-world conditions in the USA.

**Methods:** Employees of the Cleveland Clinic Health System, previously not infected with SARS-CoV-2, and working in Ohio on Dec 16, 2020, the day COVID-19 vaccination began, were included. The cumulative incidence of SARS-CoV-2 infection, over the next 5 months, was compared among those who received the vaccine and those who did not, by modeling vaccination as a time-dependent covariate in Cox proportional hazards regression analyses adjusted for the slope of the epidemic curve as a continuous time-dependent covariate.

**Results:** Of the 46866 included employees, 28223 (60%) were vaccinated by the end of the study period. The cumulative incidence of SARS-CoV-2 infection was much higher among those not vaccinated than those vaccinated. Only 15 (0.7%) of the 2154 SARS-CoV-2 infections during the study occurred among those vaccinated. After adjusting for the slope of the epidemic curve, age, and job type, vaccination was associated with a significantly reduced risk of SARS-CoV-2 infection (HR 0.03, 95% C.I. 0.02 – 0.06, p < 0.001), corresponding to a vaccine effectiveness rate of 97.1% (95% CI 94.3 – 98.5). Vaccine effectiveness was 89.2% at 7 days and 95.0% at 14 days after the first vaccine dose.

**Conclusions:** The mRNA SARS-CoV-2 vaccines are over 97% protective against COVID-19 in the working-age population, with substantial protection possibly apparent within a few days of the first dose.

**Summary:** The effectiveness of mRNA SARS-CoV-2 vaccines was evaluated among 46866 employees in an American healthcare system. After adjusting for age, job type, and the phase of the epidemic, vaccination was 97.1% effective in preventing COVID-19 among the fully vaccinated, and 89.2% protective within 7 days of the first dose.

## INTRODUCTION

Clinical trials of the mRNA vaccines against Severe Acute Respiratory Syndrome (SARS) – associated Coronavirus-2 (SARS-CoV-2) infection have found the vaccines to be highly efficacious [1,2]. One of these vaccines was also shown to have similar effectiveness in large studies conducted in Israel [3,4]. Despite these spectacular findings, vaccination efforts in the USA have been hampered by vaccine hesitancy among a substantial portion of the population.

Those reluctant to accept the vaccine might be gradually persuaded to change their minds if multiple studies show a high degree of effectiveness and safety in real-world situations within the USA. Two recent studies have shown that most healthcare personnel (HCP) in a healthcare institution were either unvaccinated or partially vaccinated when they were diagnosed with SARS-CoV-2 [5], and that the numbers of HCP infected with SARS-CoV-2 decreased with increasing days after vaccination [6], suggesting that the vaccines were very effective in preventing SARS-CoV-2 infection. Additional studies that demonstrate a high level of vaccine effectiveness would confirm the suggestion from these studies that the vaccine is highly effective.

Any study on vaccine effectiveness requires reasonably complete information about vaccination status in a population and the occurrence of COVID-19 in the same population. Since the onset of the pandemic, protecting personnel from COVID-19 has been critical for the functioning of our facilities, and it has been important for us to keep track of which employees have been afflicted by the disease and when, and which of them received the vaccine and when. The availability or such data within our Occupational Health database provided us with an opportunity to measure vaccine effectiveness within our institution.

The purpose of this study was to evaluate the effectiveness of the mRNA SARS-CoV-2 vaccines among HCP in a large American healthcare system.

## METHODS

This was a retrospective cohort study conducted at the Cleveland Clinic Health System in Ohio. PCR testing for SARS-CoV-2 was begun at Cleveland Clinic on March 12, 2020, with a streamlined process set up for testing healthcare personnel (HCP), and management of those that were found to be positive. All HCP who tested positive were interviewed by Occupational Health, and the questionnaire included the date of onset of symptoms. Shortly after approval of the first mRNA vaccine by the FDA, the institution started vaccinating its employees on December 16, 2020, with either the BNT162b2 mRNA (Pfizer-BioNTech) or the mRNA-1273 (Moderna) vaccine. A standardized process was implemented whereby HCP were scheduled for their second vaccine dose 28 days after the first, regardless of which vaccine was administered.

The study was approved by the Cleveland Clinic Institutional Review Board (IRB no. 21-300). A waiver of informed consent and waiver of HIPAA authorization were approved to allow access to PHI by the research team, with the understanding that sharing or releasing identifiable data to anyone other than the study team was not permitted without additional IRB approval.

### Screening, inclusion, and exclusion criteria

All employees of the Cleveland Clinic Health System, who were working in Ohio and were in employment, on Dec 16, 2020, were included. Those previously infected with COVID-19 (anyone who tested positive for SARS-CoV-2 at least once prior to the vaccine rollout date of Dec 16, 2020) were excluded.

### Definitions

SARS-CoV-2 infection was defined as a positive nucleic acid amplification test. When available, the date of onset of symptoms was considered the date of infection, and the date of positive test when not. A person was considered vaccinated 14 days after receipt of the second dose of the vaccine (which would correspond, in most cases, to 42 days after receipt of the first dose). Seven subjects who had been vaccinated earlier as participants in clinical trials were considered vaccinated throughout the duration of the study.

### Covariates

Job location was categorized into one of the following: Cleveland Clinic Main Campus, regional hospital (within Ohio), ambulatory center, administrative center, or remote location. Subjects were categorized into patient-facing or non-patient facing job types. Subjects were categorized into one of the following job categories: professional staff, residents/fellows, advance practice practitioners, nursing, pharmacy, clinical support, research, administration, and administration support.

### Outcome

The study outcome was time to SARS-CoV-2 infection, which was the number of days from December 16, 2020 (vaccine initiation date) to SARS-CoV-2 infection, the latter being defined as a positive nucleic acid amplification test for SARS-CoV-2 on or after December 16, 2020. HCP that had not developed a SARS-CoV-2 infection were censored at the end of the study follow-up period (May 15, 2021). Since the health system never had a requirement for asymptomatic employee test screening, most positive tests would have been associated with symptoms suspicious for COVID-19. A small proportion would have been asymptomatic infections identified because of pre-operative or pre-procedural screening.

### Statistical analysis

Vaccination (defined as having occurred 14 days after receipt of the second dose of the vaccine) was treated as a time-dependent covariate. A Simon-Makuch hazard plot [7] was created to compare the cumulative incidence of SARS-COV-2 infection among those not vaccinated (either chose not to be vaccinated or had not yet become vaccinated), and those who were vaccinated. A Cox proportional hazards model was fitted for time to SARS-CoV-2 infection against vaccination (as a time-dependent covariate whose value changed on the date the subject was considered vaccinated). The model was adjusted for age, job type, and for the phase of the epidemic (by including the slope of the epidemic curve as a continuously varying time-dependent covariate) [8,9]. The epidemic curve was created by plotting the daily proportions of all SARS-CoV-2 nucleic acid amplification tests done at Cleveland Clinic that were positive. Hazard ratios for vaccination from this model were used to calculate vaccine effectiveness (as a percentage), using the formula: vaccine effectiveness = (1 – adjusted HR) × 100. In the base case, SARS-CoV-2 infection that occurred before 14 days had elapsed since receipt of the second vaccine dose was attributed to the unvaccinated group. In order to estimate time to vaccine protection, vaccine effectiveness was measured when the duration required since receipt of vaccine for a person to be considered vaccinated was varied, starting from the day of receipt of the first dose up to 42 days later. Analyses were performed by N.K.S. and A.S.N., using the *survival* package [9] and R version 4.0.5 [10].

## RESULTS

Of 52238 employees who met inclusion criteria, 5372 were excluded because they were previously infected with COVID-19. The remaining 46866 HCP were included in the study.

### Baseline characteristics

By the end of the study, 28223 subjects (60%) were considered vaccinated (14 days or more beyond receipt of the second vaccine dose). Those vaccinated were significantly older (mean ± SD age; 44 ± 13 vs. 40 ± 13, p <0.001), and included a significantly higher proportion with patient-facing jobs (53% vs. 48%, p <0.001). Table 1 shows the characteristics of subjects grouped by whether or not they were vaccinated by the end of the study. Of those vaccinated, 63% received the Moderna vaccine. When vaccination was begun, the epidemic in Ohio was at the peak of its third wave (Figure 1).

**Table 1.** Study Subject Characteristics.

| Characteristic | Vaccinated<br>(N = 28223) | Non-vaccinated<br>(N = 18643) | P Value |
| --- | --- | --- | --- |
| Age, y, mean ± SD | 44±13 | 40±13 | <0.001 |
| Patient-facing job | 14909 (53) | 8935 (48) | <0.001 |
| Job location |  |  | <0.001 |
| Cleveland Clinic Main Campus | 12191 (43) | 6457 (35) |  |
| Regional hospitals | 8542 (30) | 6709 (36) |  |
| Ambulatory centers | 4377 (16) | 2971 (16) |  |
| Administrative centers | 2437 (9) | 1783 (10) |  |
| Remote location | 676 (2) | 723 (4) |  |
| Job category |  |  | <0.001 |
| Professional staff | 3134 (11) | 528 (3) |  |
| Residents and fellows | 1398 (5) | 200 (1) |  |
| Advanced practice practitioners | 1688 (6) | 938 (5) |  |
| Nursing | 7300 (26) | 5247 (28) |  |
| Pharmacy | 826 (3) | 415 (2) |  |
| Clinical support | 3034 (11) | 3350 (18) |  |
| Administration | 2327 (8) | 1011 (5) |  |
| Administration support | 7728 (27) | 6579 (35) |  |
| Research | 718 (3) | 444 (2) |  |

Data are presented as no. (%) unless otherwise indicated

**Figure 1.**
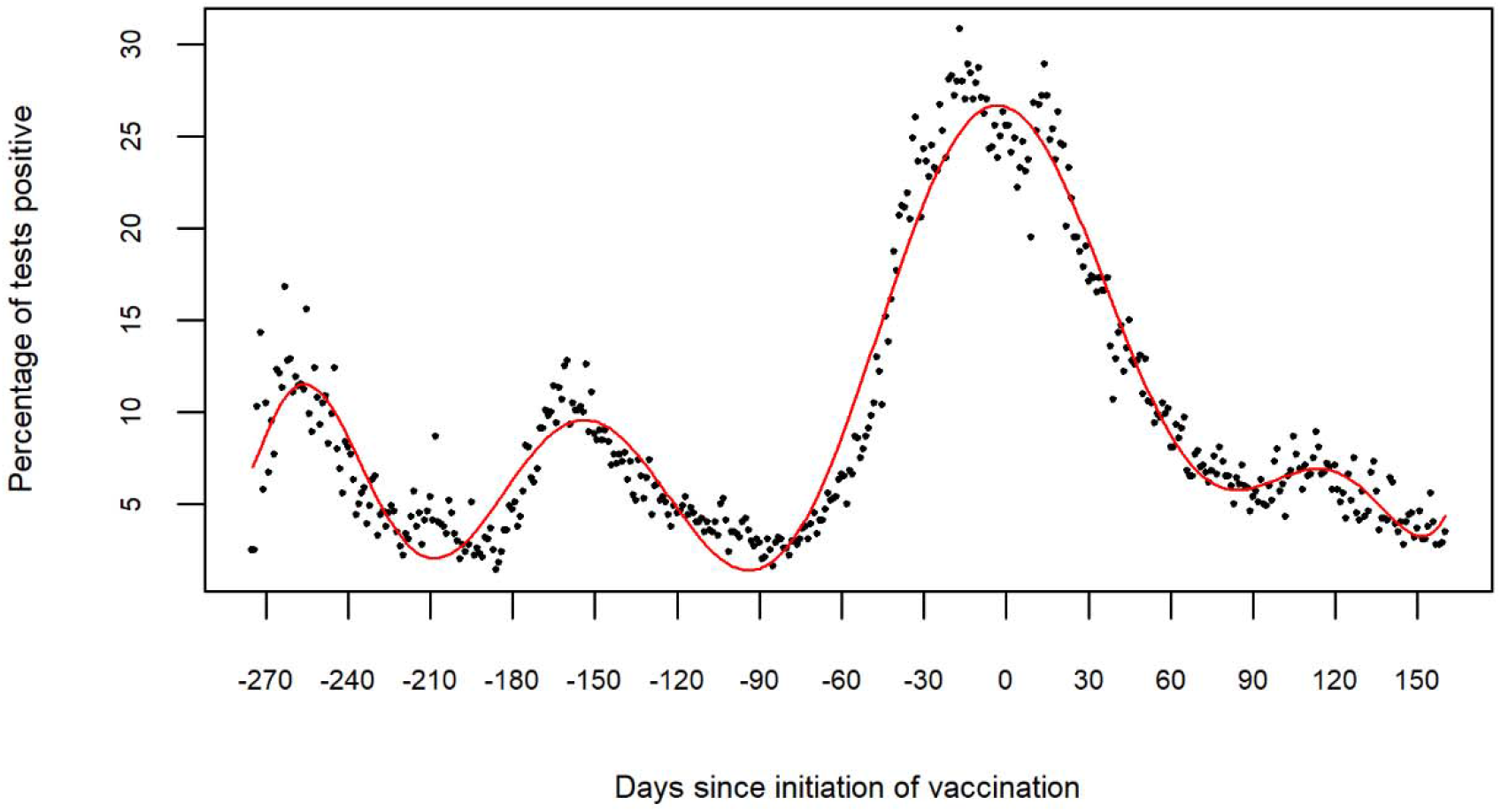
COVID-19 epidemic curve before and after vaccine rollout. Day zero on the x-axis represents the day vaccination began at the health system. Points on the scatter plot represent the proportion of tests that were positive on any given day. The colored line represents a fitted polynomial curve.

### Cumulative incidence of COVID-19

Figure 2 is a Simon-Makuch plot showing that SARS-CoV-2 infections occurred almost exclusively in subjects who were not vaccinated. The cumulative incidence of SARS-CoV-2 infection among those vaccinated was almost zero regardless of whether subjects received the Pfizer-BioNTech or the Moderna vaccine. By the end of the study period, every HCP in our health system had ample opportunity to receive at least one dose of the vaccine, and those who had not yet received at least one dose of the vaccine in almost all cases were those who chose to remain unvaccinated. If those not vaccinated were separated into those who chose not to get vaccinated (i.e. those who did not receive a single dose of the vaccine throughout the course of the study) and those who were willing to take the vaccine but were not yet considered vaccinated (i.e. either they had not yet received the vaccine or it had not yet been 14 days since receiving the second vaccine dose), those not yet considered vaccinated had a lower cumulative incidence of SARS-CoV-2 infection than those who chose not to get vaccinated (Figure 3). Of the 2154 SARS-CoV-2 infections during the study period, 1344 (62.4%) occurred among those who chose not to get vaccinated, 795 (36.9%) occurred while the affected subject was not yet considered vaccinated, and only 15 (0.7%) occurred in those who were considered vaccinated.

**Figure 2.**
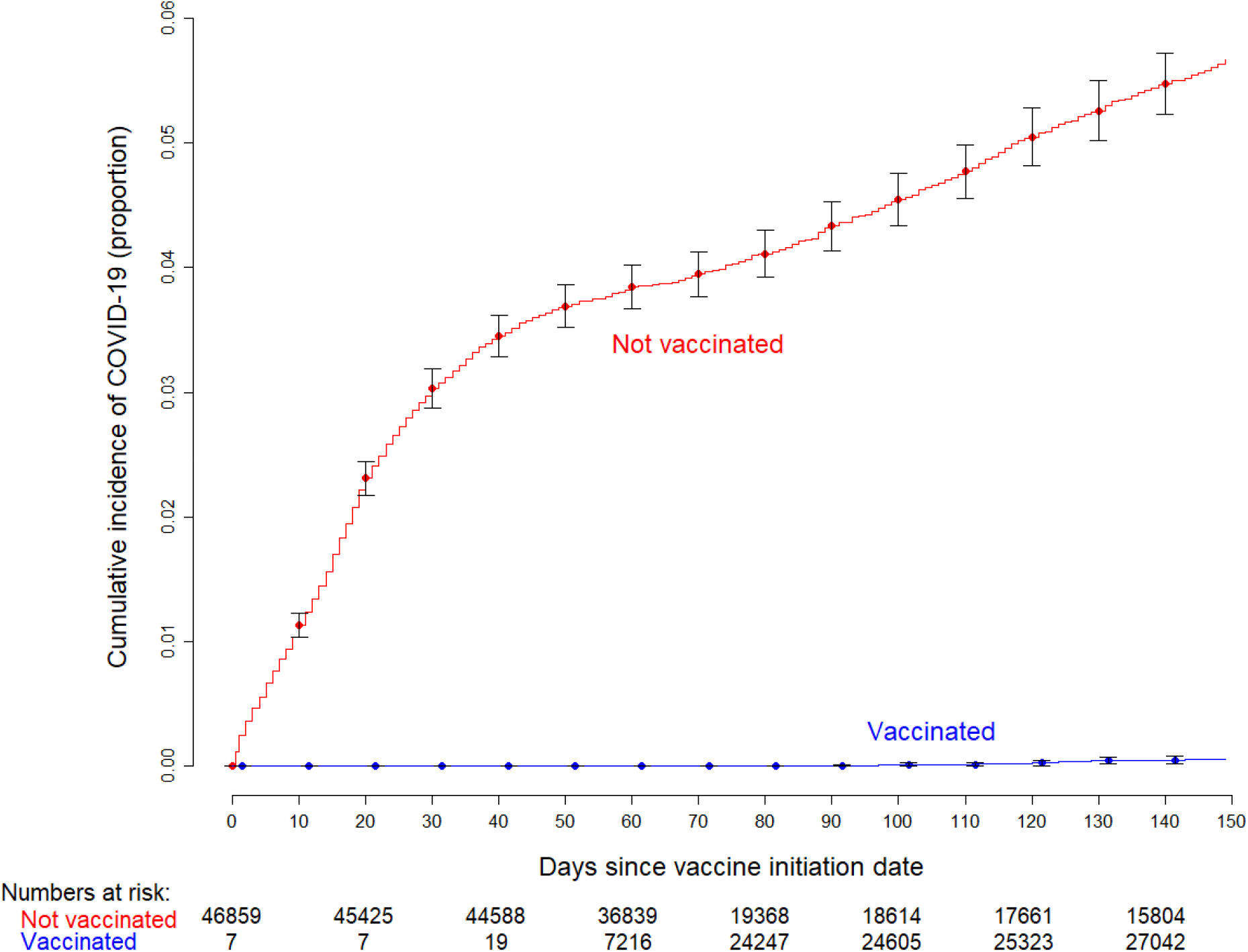
Simon-Makuch plot showing the cumulative incidence of COVID-19 among vaccinated and not vaccinated subjects. The curve for the not vaccinated group includes data for those who never received the vaccine at any point during the study and those who were waiting to become vaccinated. Seven subjects who had been vaccinated earlier as participants in clinical trials were considered vaccinated throughout the duration of the study. Twelve subjects who received their first dose in the first week of the vaccination campaign managed to get their second dose three weeks later, and were thus considered vaccinated earlier than 42 days since the start of the vaccination campaign.

**Figure 3.**
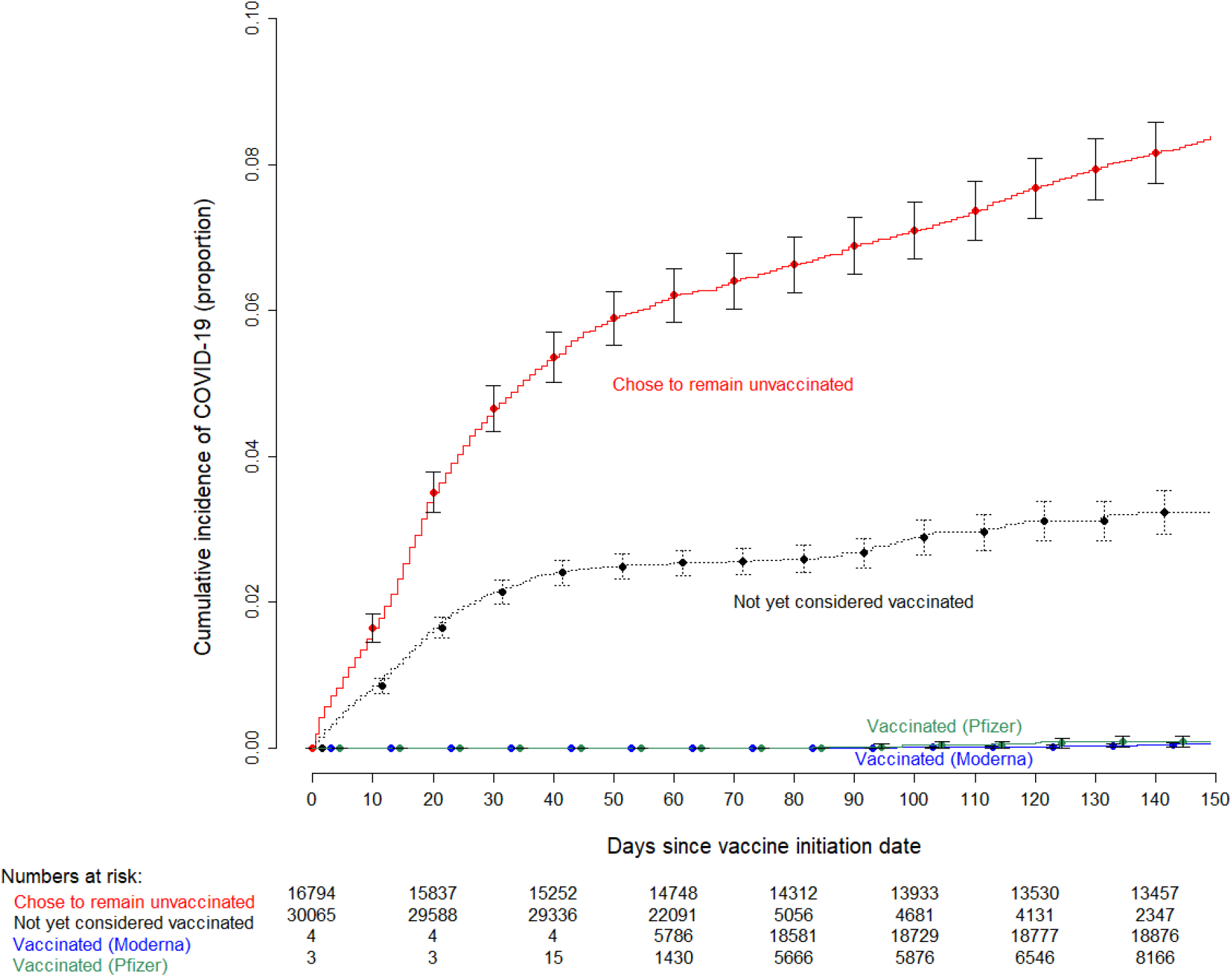
Simon-Makuch curve comparing the cumulative incidence for COVID-19 among those who remained unvaccinated, those not yet vaccinated (waiting to receive the vaccine), and those who received the Moderna or Pfizer vaccine. Seven subjects who had been vaccinated earlier as participants in clinical trials were considered vaccinated throughout the duration of the study. Twelve subjects who received their first dose in the first week of the vaccination campaign managed to get their second dose three weeks later, and were thus considered vaccinated earlier than 42 days since the start of the vaccination campaign.

### Vaccine effectiveness

In a Cox proportional hazards regression model, after adjusting for the phase of the epidemic, age, and job type, vaccination was associated with a significantly lower risk of SARS-CoV-2 infection (HR 0.03, 95% C.I. 0.02 – 0.06, p < 0.001). Estimates for unadjusted and adjusted hazard ratios for the variables included in the model are shown in Table 2.

**Table 2.** Unadjusted and Adjusted Hazard Ratios for Factors associated with SARS-CoV-2 Infection in Cox Proportional Hazards Regression Models.

| Variable | Unadjusted HR (95% CI) | Adjusted HR (95% CI) |
| --- | --- | --- |
| Vaccination<sup>1</sup> | 0.02 (0.01 – 0.05) | 0.03 (0.02 – 0.06) |
| Slope of the epidemic curve<sup>2</sup> | 24.15 (14.05 – 41.50) | 0.91 (0.66 – 1.26) |
| Age (per year) | 0.982 (0.979 – 0.985) | 0.984 (0.981 – 0.987) |
| Patient-facing job | 1.13 (1.04 – 1.23) | 1.09 (0.99 – 1.18) |

<sup>1</sup>Defined as having occurred 14 days after receipt of the second dose of a mRNA SARS-CoV-2 vaccine. This was modeled as a time-dependent covariate.
<sup>2</sup>The slope of the epidemic curve was the first derivative of the epidemic curve, which was a polynomial curve fitted to the daily proportion of all SARS-CoV-2 nucleic acid amplification tests at Cleveland Clinic that were positive. The slope of the epidemic curve was modeled as a continuous time-dependent covariate.

Based on the above multivariable model, vaccine effectiveness for protection from SARS-CoV-2 infection was 97.1% (95% CI 94.3 – 98.5).

### Time to vaccine protection

There appeared to be substantial protection from SARS-CoV-2 infection within a few days of the first dose of an mRNA vaccine. Vaccine effectiveness increased as the definition of vaccinated required more days to have elapsed after the first dose of the vaccine (Figure 4), and was already very high within a few days of the first vaccine dose. Vaccine effectiveness was 89.2% at 7 days and 95.0% by 14 days after receipt of the first dose of the vaccine. Table 3 displays the findings of vaccine effectiveness if a person was considered vaccinated at 7, 10, 14, 21, and 28 days, after the first vaccine dose.

**Figure 4.**
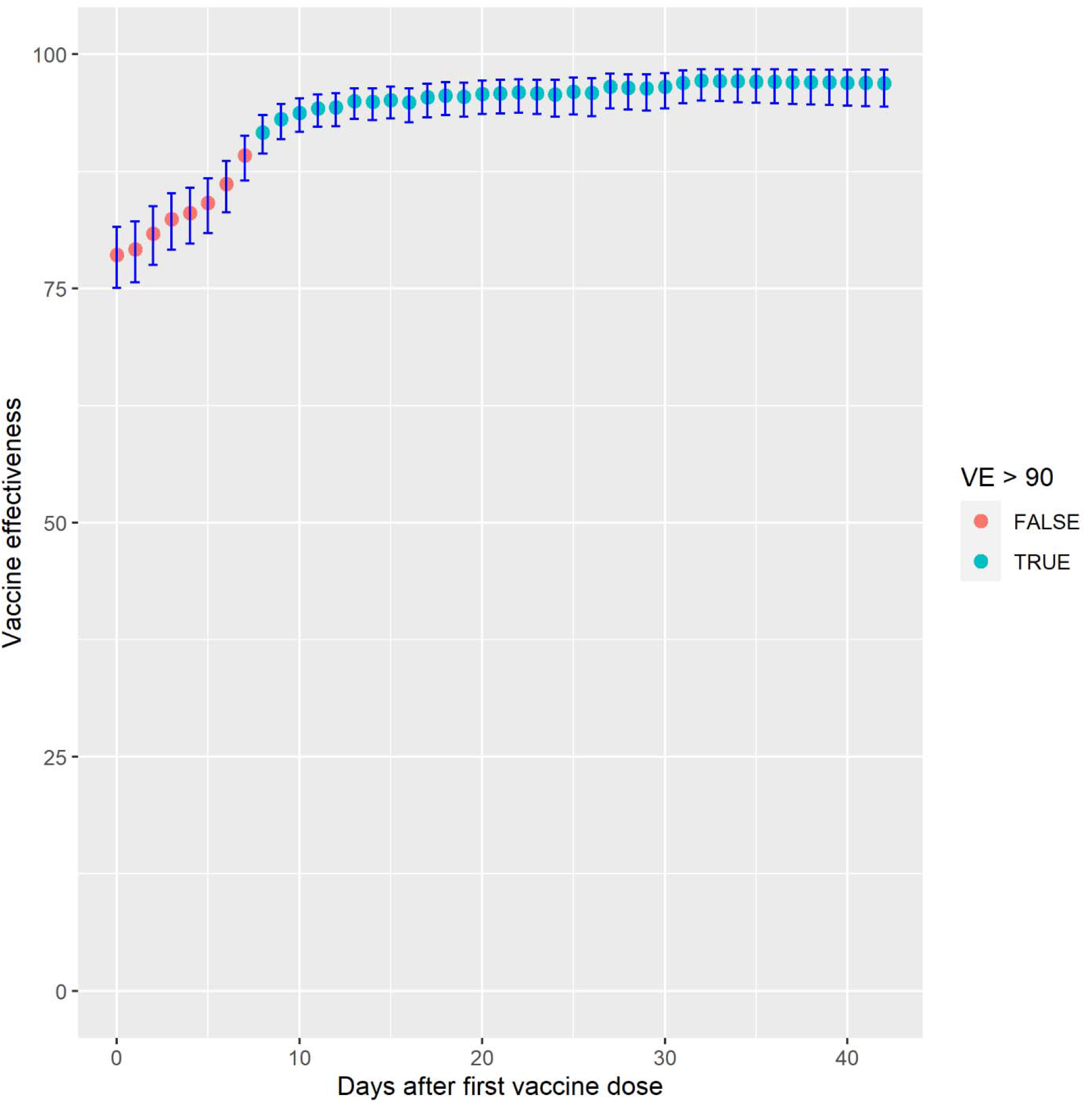
Estimates of vaccine effectiveness at different days after receipt of the first dose of the vaccine. Points represent point estimates and the error bars represent 95% confidence intervals. Vaccine effectiveness estimates above 90% are colored blue, while those 90% or less are colored red.

**Table 3.** Vaccine Effectiveness when the Definition of Vaccination is Varied by Days after Receipt of First Dose of the Vaccine.

| Number of Days after First Dose | Vaccine Effectiveness<br>(%) | 95% CI |
| --- | --- | --- |
| 7 days | 89.2 | 86.5 – 91.3 |
| 10 days | 93.8 | 91.7 – 95.3 |
| 14 days | 95.0 | 93.0 – 96.4 |
| 21 days | 95.8 | 93.7 – 97.3 |
| 28 days | 96.5 | 94.1 – 97.9 |

## DISCUSSION

This study found that the mRNA SARS-CoV-2 vaccines were 97.1% effective in protecting against SARS-CoV-2 infection, an effectiveness that was similar to vaccine efficacy noted in previously published clinical trials [1,2], and in large vaccine effectiveness studies in Israel [3,4].

An interesting finding in our study is that the cumulative incidence of SARS-CoV-2 infection among those waiting to become vaccinated, although higher than those vaccinated, was lower than those who remained unvaccinated. The latter would have largely included individuals who opted not to receive the vaccine despite ample opportunity to do so. The difference in cumulative incidence of the infection suggests different risk avoidance behaviors in those willing and unwilling to take the vaccine.

In addition to confirming a very high vaccine effectiveness rate in a real-world setting, this study also suggests that the mRNA vaccines may provide substantial protection within a few days of administration of the first dose of the vaccine. One must be careful not to interpret this to mean that the second dose is not necessary. This study was not designed to examine the necessity of a second dose, and most persons in this study (96% of those eligible) received two doses of the vaccine. This study did find, however, that the protective effect of a vaccine was apparent within a few days of individuals receiving their first dose of a two-dose series. Whether this was because the vaccine provides protection much earlier than is widely believed or whether this might have been from continued risk avoidance measures by those who chose to be vaccinated, will have to be clarified in future studies.

This study’s strengths are that it was large enough to detect meaningful differences across comparison groups, and that the phase of the epidemic curve was adjusted for. As expected, the study found that the risk of SARS-CoV-2 infection was very strongly associated with the phase of the epidemic, and increased as the slope of the epidemic curve increased. Accordingly, in a vaccinated individual, the risk of acquisition of infection over time would depend on the phase of the epidemic at the time the person received the vaccine. Our multivariable models were adjusted for this confounder.

A limitation of our study is that the absence of a program of systematic testing of asymptomatic individuals would have resulted in most asymptomatic infections being missed. This study therefore cannot exclude the possibility that asymptomatic infections might have occurred despite vaccination. This is not a crippling limitation, however, as asymptomatic patients would have been missed in both the unvaccinated and vaccinated groups. It is also theoretically possible that vaccinated persons were less likely to be tested than unvaccinated ones when suspicious symptoms arose. This is probably less of a limitation among our HCP than it might have been in other populations because of easy access to testing (no co-payment required and streamlined testing process for HCP), and the expectation of HCP being exposed to the disease during the course of their work, which would have increased the likelihood of testing for COVID-19 if suspicious symptoms arose. Certain groups were not represented in our study, including children and the elderly, and the number of immunocompromised patients would have been expected to have been small. Caution is advised in in extrapolating the findings of the study to these groups.

Our study’s findings add to the growing literature on the effectiveness of the mRNA SARS-CoV-2 vaccines. These findings are consistent with other studies that have examined this question using different approaches at different sites in the USA [5,6,11]. The high rates of vaccine effectiveness even when assuming a person vaccinated within a few days of the first dose, suggests the possibility that the vaccine may be protective well before the second dose is administered.

In conclusion, the Pfizer-BioNTech and Moderna vaccines are highly effective in protecting individuals among the working-age population from SARS-CoV-2 infection in real-world conditions in the USA, the vaccines may provide substantial protection long before receipt of the second dose of the vaccine, and SARS-CoV-2 infection in this population at this time is occurring almost exclusively among those not vaccinated.

## Data Availability

De-identified individual level data and code to reproduce the results are available in a public data repository.

https://osf.io/r9n5u/

## Notes

### Competing Interest Statement

The authors have declared no competing interest.

### Funding Statement

No external funding.

